# From Necessity to Preference: A Study of Predictors Influencing Elective Caesarean Section in Rwanda

**DOI:** 10.1101/2024.05.01.24306711

**Authors:** Munawar Harun Koray, Theophile Dushimirimana, Tanya Curry, Katia Olaro Adupo, Alfred Pie Faabie, Damien Punguyire

## Abstract

**Background:** Caesarean section is an important obstetric intervention that saves the lives of mother and newborn babies. However, its increase is of global public health concern. Despite tremendous reduction in maternal and newborn morbidity and mortality, Rwanda has shown a very high incidence of CS among mothers in recent years. Therefore, this study investigated the predictors of patient-initiated elective CS in Rwanda.

**Methods:** In this cross-sectional study, we used nationally representative data from Rwanda Demographic and Health Survey 2019/20. A total of 6,167 females were included in this study. Chi-square test was used to test association between the type of caesarean section and demographic characteristics. Both binary and multivariate regression analysis were performed to assess the predictors of elective caesarean section at a p-value ≤ 0.05 and 95% confidence interval. Model fitness was rigorously conducted to ensure validity and reliability of study findings. The data was analysed using STATA version 14 SE.

**Results:** The rate of CS among women who delivered (6,167) in the last five years preceding the survey was 1,015 (16.46%). Among the women who underwent CS, 36.6% opted for elective CS. Our findings showed that women aged 30 – 39 years were more likely to opt for elective CS [aOR: 3.130, 95%CI:1.969 - 4.978] compared to those aged 29 years or below. Women living in rural areas were less likely to opt for elective CS in the binary regression model [cOR: 0.587, 95%CI: 0.448 – 0.768]. Also, mothers who received ANC assistance by nurse/midwife were 40% less likely [aOR: 0.529, 95%CI: 0.349 – 0.803] to undergo elective CS, than those assisted by doctors.

**Conclusion:** The rate of elective CS is very high among mothers in Rwanda. By using the 2019/20 RDHS data, the study found the key predictors behind the high rate of CS in Rwanda. These predictors should be deeply considered in developing comprehensive measures and policies to mitigate the unnecessary CS in Rwanda, which has detrimental impact on maternal and newborn outcome.

## Introduction Background

Caesarean Section (CS) is the most common surgical procedure performed on women around the world (1). It was introduced in the late Nineteenth century as an obstetric intervention to save lives of mothers and their newborn babies from pregnancy- and childbirth-related complications during labor (2,3). This procedure is primarily recommended for cases such as placenta praevia, fetal malposition, uncontrolled hypertension or diabetes in the mother, multiple pregnancies, or previous CS (4,5). Although CS is a live-saving intervention, studies have shown that unwarranted CS may have increased risk of maternal, neonatal and infant mortality and morbidities (1,6–8).

A Canadian study involving a 14-year follow-up of over two million women who underwent CS with low-risk pregnancy in their first CS were three times more to die or suffer serious complications, including infections, blood clots, and heart attack, compared to their counterparts who delivered vaginally (9). Also, children born through CS are found to have a higher risk of developing respiratory tract infections (RTIs), obesity, and asthma, compared with children born vaginally (1). Despite these concerns, the global incidence of Caesarean section (CS) has surged in recent years, prompting an extensive debate and research into its medical, social, and economic drivers.

Data from 2010 to 2018 across 154 countries shows an increase in CS rates, where more than 1 in 5 (21.1%) of all childbirths are born through CS, with averages ranging from 5% to 42.8% in sub-Saharan Africa (SSA) and Latin America and the Caribbean, respectively (10,11). It is projected that by 2030 28.5% of women worldwide will give birth through CS, averaging from 7.1% in SSA to 25.7% in Europe and more than 60% in Eastern Asia (10,12). Rwanda is one of the countries in SSA region with very high incidence of CS, increasing from 2.2% in 2000 to 15.6% in 2019 (13).

The increasing risk of complications related to CS prompted the WHO to recommend CS rates in countries not to exceed 10-15% (14). Despite this recommendation, more women are undergoing CS at undesirable rates, with studies citing reasons such as perceived safety of CS, “pain-free” mode of delivery, partner’s preference for CS, and bad experience with previous vaginal delivery (15–17). Other factors such as maternal age, socio-economic status, occupation, educational levels, number of antenatal care (ANC) visits and birth order have been linked to increase risk of CS (3,13,17).

In recent years, Rwanda has achieved a tremendous reduction in maternal and newborn mortality, with a 79% decline in maternal mortality ratio between 2000 and 2017 (18). These reductions have been largely attributed to improved access and use of skilled delivery (13). However, there is a risk of setback in this gain if unnecessary CS are not acted upon to reduce the rate of CS in Rwanda. Previous studies have sort to identify the determinants of CS in Rwanda, as steps to reversing the unnecessary increase in CS in the country (13,19). However, these studies were limited to the broader scope of CS, without recourse to the actual prompt for CS; either emergency or patient-initiated elective CS. The occurrence of patient-initiated elective CS highlights the growing trend of mothers opting for their preferred method of childbirth (20). Therefore, this study investigated the predictors of patient-initiated elective CS in Rwanda. The study used a nationally-representative survey, the 2019-20 Rwanda Demographic Health Survey (RDHS), modelling the data to increase its validity and reliability.

## Objective of the Study

The objective of this study is to investigate the predictors of elective CS in Rwanda using a sound modelled data from RDHS

## Methods

### Study Design

The data for this study was obtained from the 2019-20 Rwanda Demographic and Health Survey (RDHS). The RDHS is a nationally representative cross-sectional survey conducted by the National Institute of Statistics of Rwanda (NISR), in collaboration with the Ministry of Health (MOH) of Rwanda, with technical from ICF through the Demographic Health Program (DHS). This survey offers extensive national data that spans the entire spectrum of public health, with a particular emphasis on maternal and child health. The data utilized for this study is accessible online at: https://www.dhsprogram.com/data/available-datasets.cfm (21).

The 2019–20 RDHS used a two-stage sample design and was designed to enable estimates of select limited variables for each of Rwanda’s 30 districts, five provinces, urban and rural areas, and the country as a whole. In the first stage, sample sites (clusters) made up of the EAs defined for the 2012 RPHC were identified. A total of 522 clusters, 112 in urban and 388 in rural areas were chosen.

In the second stage, a systematic sample of households were selected. From June to August 2019, a household listing operation was conducted in each of the chosen EAs. Households that would be part of the survey were chosen at random from these lists. A total of 13,005 households were included in the sample, with 26 houses chosen from each sample point. The sample is not self-weighting at the national level due to the almost similar sample sizes in each district; therefore, weighting factors have been added to the data file to ensure that the results are proportionate at the national level. Women aged 15 to 49 who were either long-term residents of the chosen homes or guests who spent the night before the survey were eligible for the study. This study design is guided by the Strengthening the Reporting of Observational Studies in Epidemiology (STROBE) (22) (Supplementary material 1).

### Study setting

Rwanda is located in East Africa with a population above 13 million in 2022, leaning towards younger age groups with median age of 19 years. (23). It is border by Democratic Republic of Congo, Tanzania, Uganda, and Burundi. With a population density of 499 people per square kilometer, Rwanda ranks as the second most densely populated region in Sub-Saharan Africa (SSA) with a population density of 546 individual/km^2^ in 2021 (24). About 86% of households subscribe to Community-Based Health Insurance (CBHI) to cover healthcare costs (25). Rwanda is partitioned into 5 provinces, with 30 districts. Each district is sub-divided into enumeration areas (EA) using the 2012 Rwanda Population and Housing Census.

### Population and Sample Size

The population for this study is made up of females age 15 – 49 years, who were permanent or temporary residents of households in the previous night prior to the survey were selected. The sample included 13,005 selected households, out of which 12,951 were occupied. Interviews were successfully conducted in 12,949 of these occupied households, resulting in a response rate of 100%. The study sample was narrowed to only females who had a history of child birth (normal delivery or CS) in the last five years prior to the DHS survey. A total of 6,167 females were therefore included in this study.

**Figure 1:**
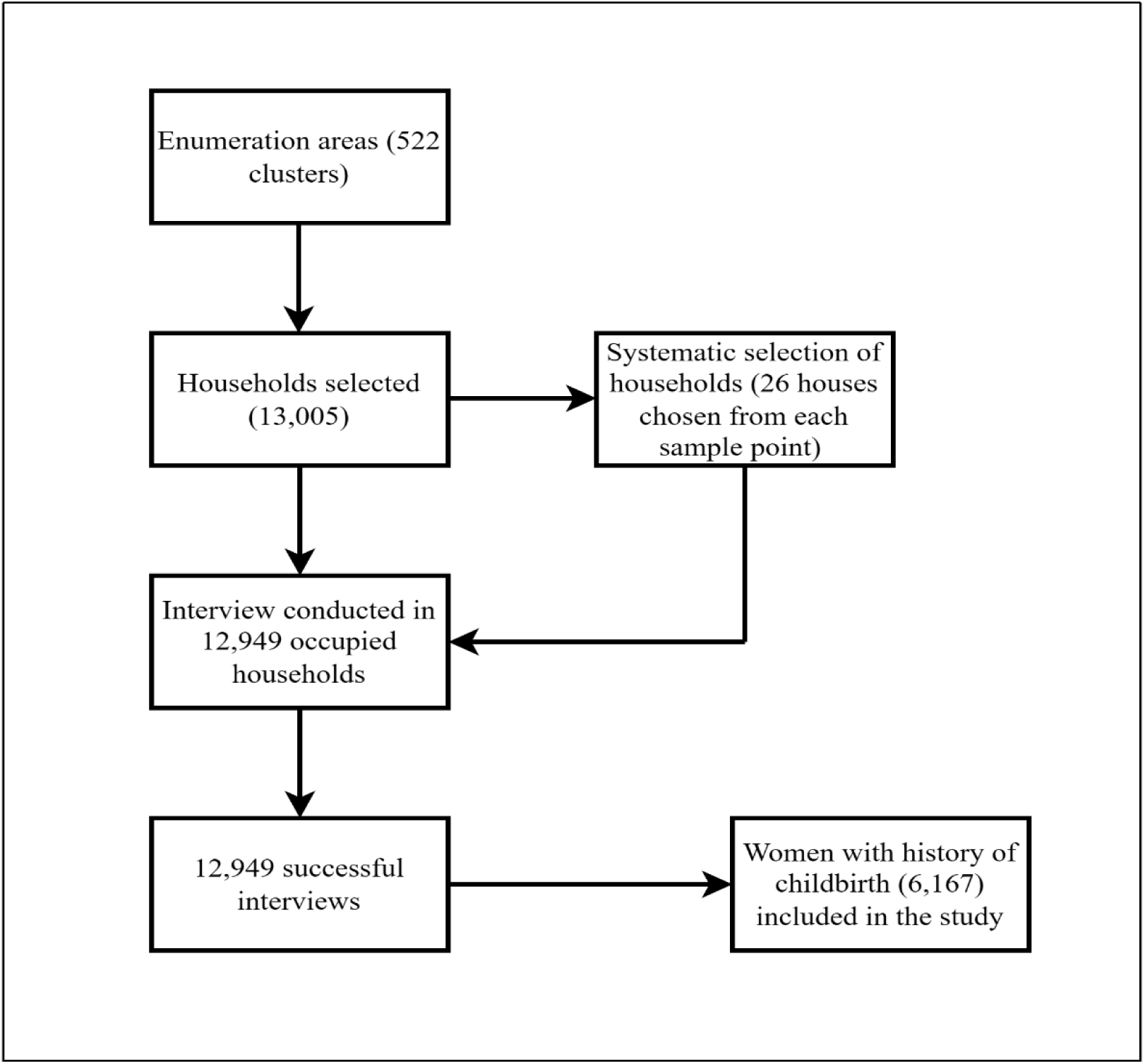
Participant selection process

### Variables

#### Outcome variable

The outcome variable for this study is CS. Respondents were asked if they underwent CS during delivery. Their responses were coded 0 “No” and 1 “Yes”. However, the variable did not include the type of CS, whether it was elective CS or emergency CS. Therefore, the type of CS was obtained by recoding the “timing on decision for caesarean section” variable in the dataset, coded 1 “decision before labor started” and 2 “decision after labor started”. It was taken that decision before labor started is “elective CS” (coded as 0) and decision after labor started is “emergency CS” (coded as 1).

#### Independent variables

The independent variables for this study were variables found to be significantly associated with CS in previous studies (26–29). These include age (recoded as: less than 20, 20 – 29, 30 – 39, 40 – 49), marital status (never married, married, cohabitation, widowed, divorced), educational status (no formal education, primary education, secondary education, higher education), religion (recoded as: Christianity, Islam, Traditional/animalist, no religion, Other), parity (recoded as: 1 – 3, 4 – 6, 7 – 9, 10 or more), occupation (recoded as: did not work, Professional/technical/managerial, clerical, agriculture, household and domestic service, Others), wealth index ( recoded as: Poorest, Poorer, Middle, Richer, Richest), place of residence (rural and urban), exposure to media (frequency of reading newspaper/magazine, frequency of listening to radio, frequency of watching television), gestational age at ANC registration (recoded as: No ANC, < 4, 4 – 5, 6 – 7, 8 or months and Don’t know), number of ANC attendance (recoded as: None, <4, 4 – 8, and 9+), person providing assistance during ANC (recoded as: Doctor, nurse/midwife, other health worker, No ANC), received 2+ tetanus injections (No and Yes), took iron tablet/syrup during pregnancy (No or Yes), took intestinal parasite drugs during pregnancy (No and Yes).

#### Data Source

The data for this study was obtained from the responses on the women questionnaire of the DHS program (https://dhsprogram.com/Methodology/Survey-Types/DHS-Questionnaires.cfm#CP_JUMP_16179). The questionnaire was used to obtain data on background characteristics (such as age, education, religion, ethnicity, occupation, wealth index, etc), maternal and child healthcare services, and partner’s background characteristics in the nation-wide survey. The data was extracted from the DHS website in a STATA format.

### Patient and Public Involvement

No patient or the public were involved in the study.

### Data Analysis

The data was analysed using STATA version 14 (StataCorp LP, College Station, Texas, USA). The study used a design-based analysis approach with weighting to adjust for the unequal probabilities employed by the DHS. This was done to reduce sample variability for particular regions or subpopulations while maximizing case representation (30). Weighting was done to cater for sampling and design bias.

The study analysis was done in three steps. In the first step, descriptive statistics was used to summarize the frequency of the types of CS and chi-square test (ꭓ^2^) was used to test the association between the independent variables and the outcome variable (type of CS). In the second step, a backward stepwise logistic regression was used to identify strong predictors of elective CS and to reduce overfitting of the regression model (31,32). Independent variables that were significant at p-value ≤ 0.25 in the chi-square test were put into the regression model (33). At each step, variables with p-value > 0.25 were eliminated until all independent variables were significant at p ≤ 0.25. the final model was used for the binary logistic regression.

In the third stage of the analysis, binary logistic (crude odds ratio[cOR]) and multivariate logistic regression (adjusted odds ratio [aOR]) were employed to identify the predictors of elective CS in Rwanda. Variables that were significant at p-value ≤ 0.05 at 95% confidence interval in the binary logistic regression model were put in the multivariate logistic regression model to control for confounders. The data did not contain missing data, thus the standard statistical methods were applied without adjustments.

### Model Diagnostics

Variance Inflation Factor (VIF) was used to assess the multicollinearity among the independent variables that were selected after the backward stepwise analysis to ensure the model validity (34). VIF values more than 5 or 10 are often considered as a show of multicollinearity (35–37). The VIF value of greater than 5 was considered collinearity (35). The goodness of fit for the model was again evaluated using Hosmer-Lemeshow test to provide a robust framework for interpreting the reliability of the results (38). Additionally, Akaike Information Criterion (AIC) was used to further examine the fitness of the final model compared to the original model (39). The final model with lowest AIC value indicates parsimonious model, confirming its reliability. Likelihood Ratio Test (LR-test) was also used to compare the goodness of fit between the final model nested into the original model (40). A non-significant p-value indicates that the final model presents a better fit for the data.

## Results

### Caesarean Section Rates

The rate of CS among women who delivered (6,167) in the last five years preceding the survey was 1,015 (16.46%). Among the women who underwent CS, 36.6% opted for elective CS (figure 2).

**Figure 2:**
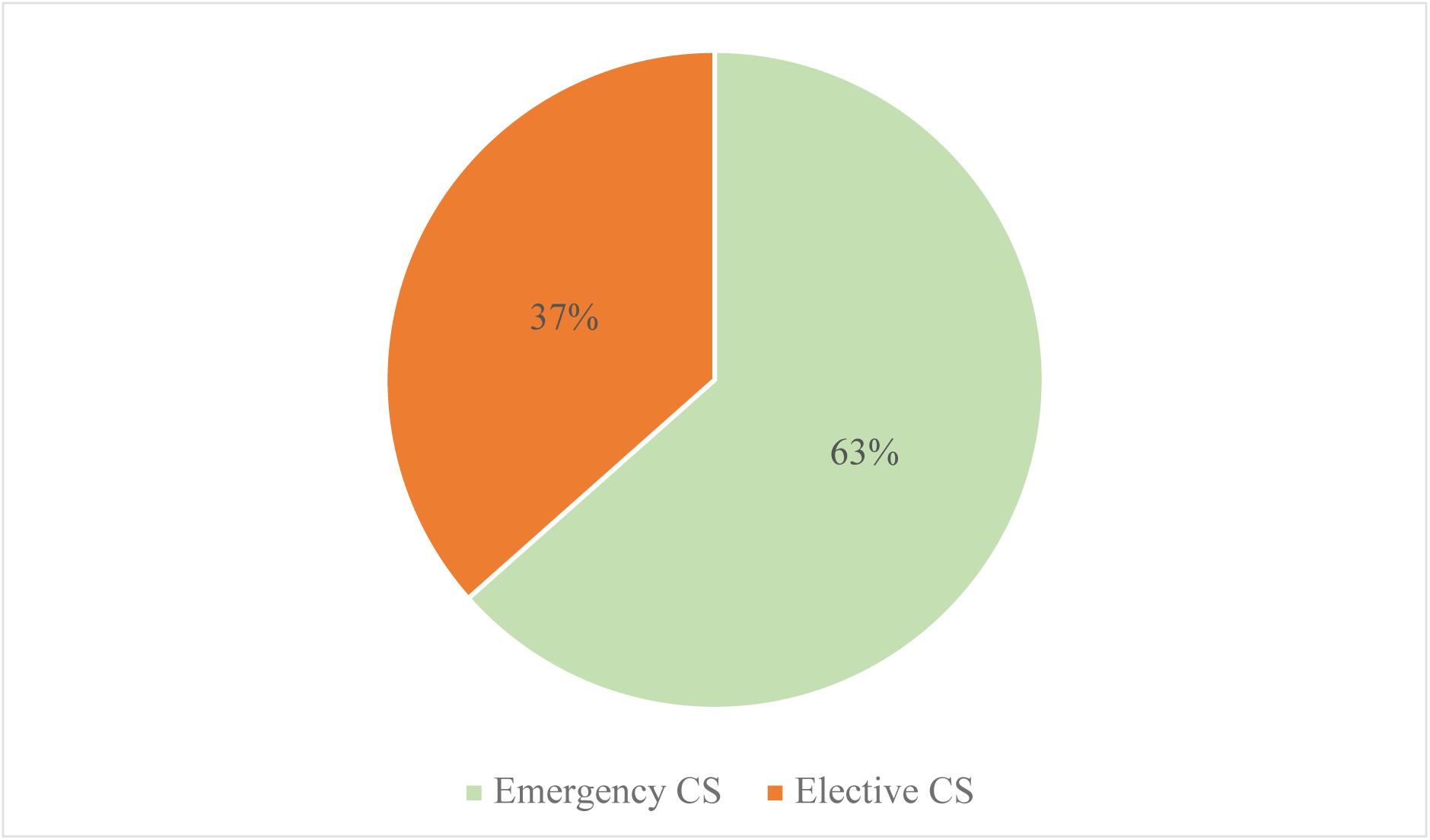
Frequency of type of CS

Table 1 presents the socio-demographic characteristics and the distribution of Cesarean section (CS) types. The data reveals a significant association between age and CS (p < 0.0001), with the highest rate of elective CS (44.7%) observed among women aged 30 to 39 years. Education level also shows a significant association with CS (p = 0.035), with mothers with higher education levels having the highest rate of elective CS (47.73%). Factors such as parity (p < 0.0001), wealth index (p = 0.018), and place of residence (p < 0.0001) were similarly linked to CS rates. Specifically, mothers in the richest wealth index category (43.07%) and those residing in urban areas (45.0%) exhibited the highest rates of elective CS. Additionally, the type of professional providing assistance during antenatal care (ANC) was significantly associated with CS (p < 0.0001), with 55.4% of mothers attended by a doctor opting for elective CS.

**Table 1:**
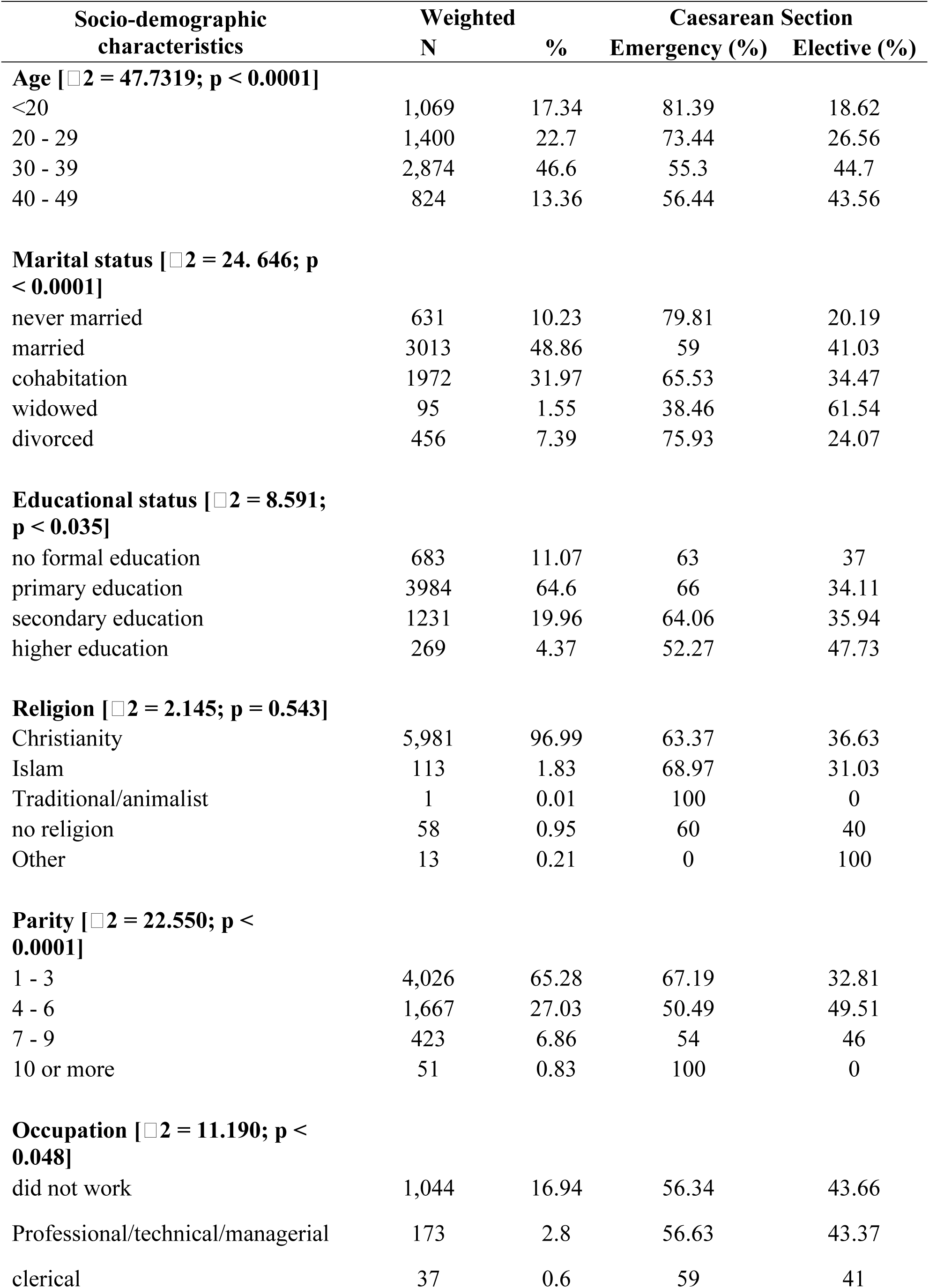
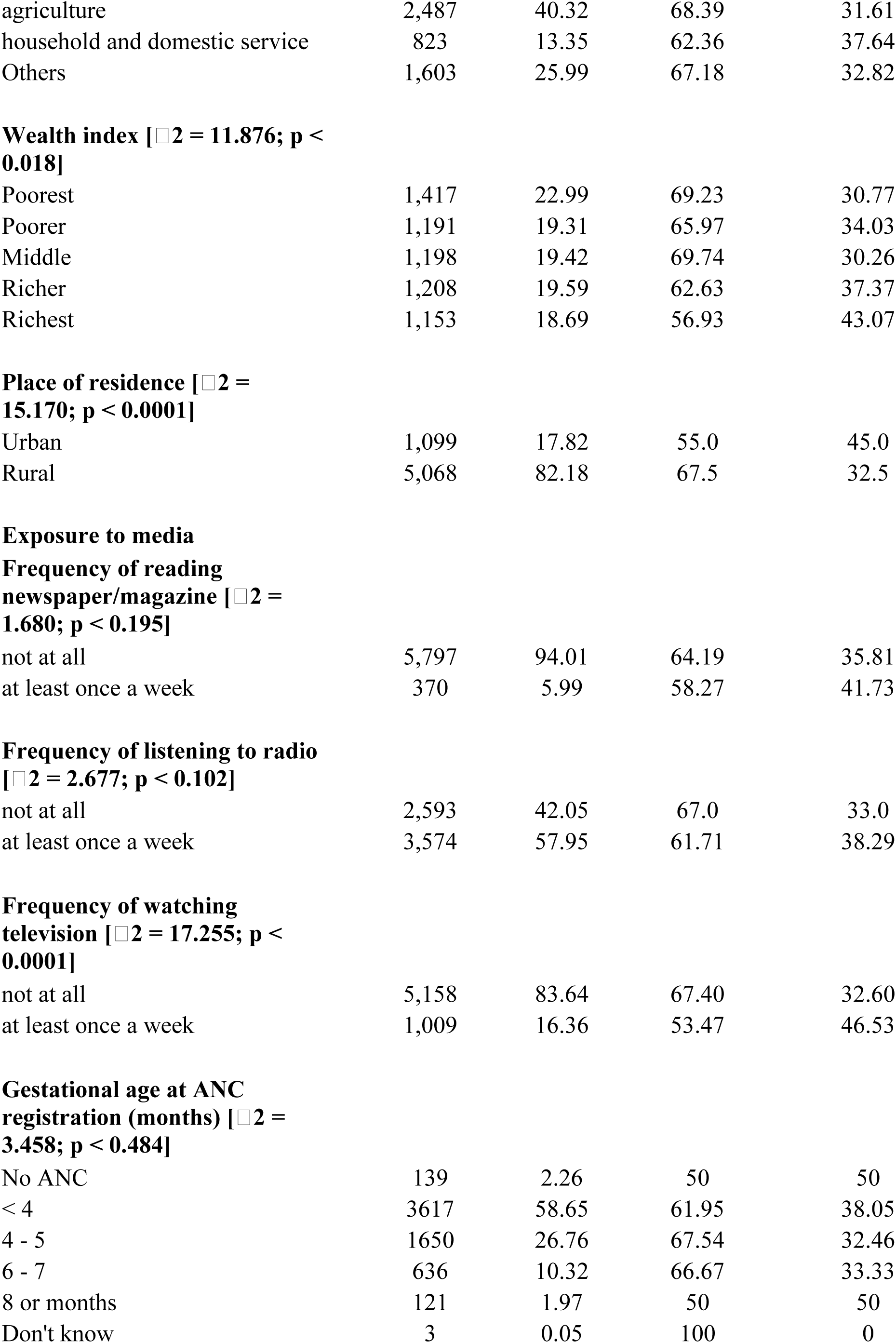
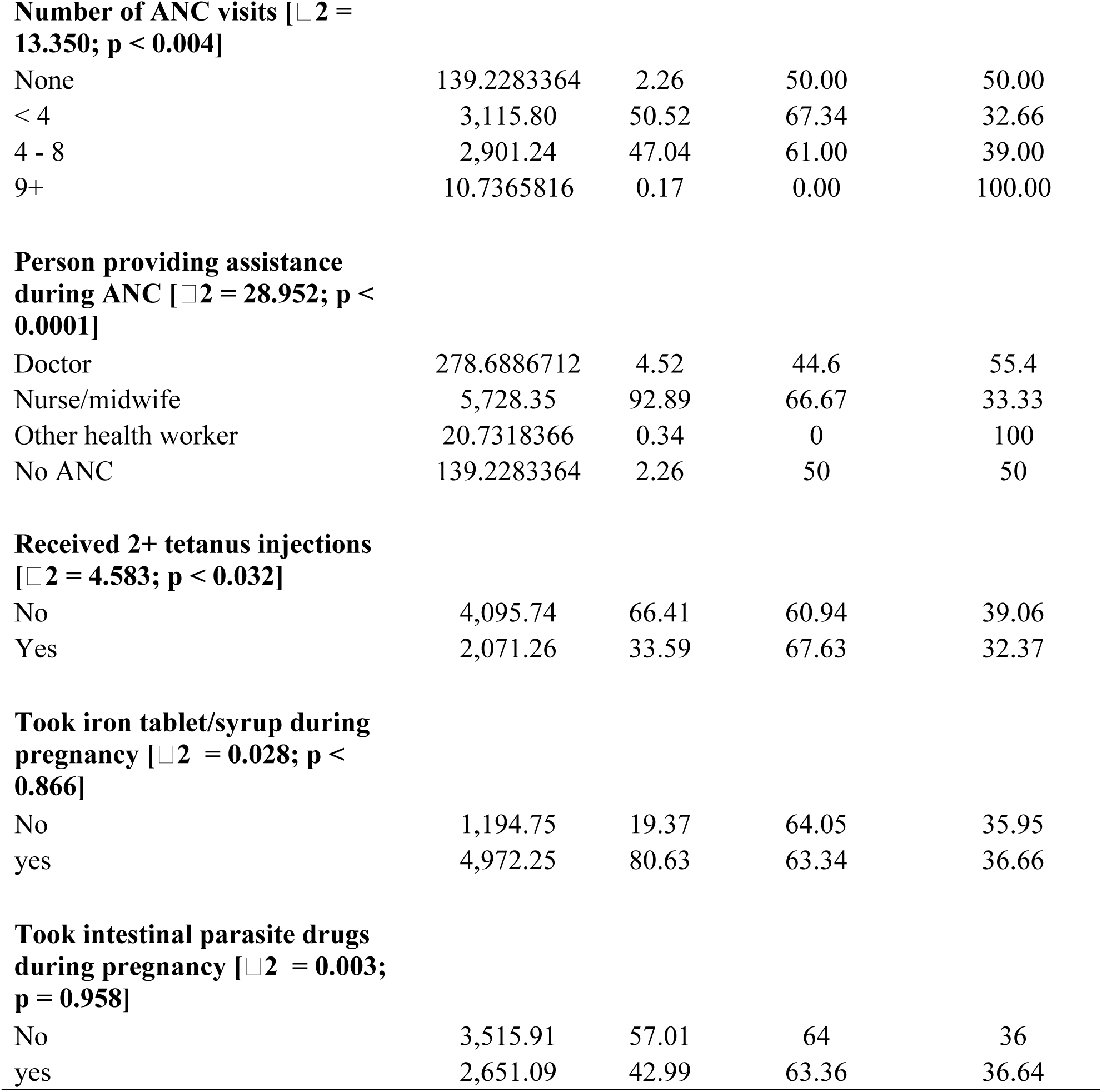
Socio-demographic Characteristics and Distribution of Caesarean Section.

| Socio-demographic characteristics | Weighted |  | Caesarean Section |  |
| --- | --- | --- | --- | --- |
|  | N | % | Emergency (%) | Elective (%) |
| **Age [$\chi^2 = 47.7319$; $p < 0.0001$]** |  |  |  |  |
| <20 | 1,069 | 17.34 | 81.39 | 18.62 |
| 20 - 29 | 1,400 | 22.7 | 73.44 | 26.56 |
| 30 - 39 | 2,874 | 46.6 | 55.3 | 44.7 |
| 40 - 49 | 824 | 13.36 | 56.44 | 43.56 |
| **Marital status [$\chi^2 = 24.646$; $p < 0.0001$]** |  |  |  |  |
| never married | 631 | 10.23 | 79.81 | 20.19 |
| married | 3013 | 48.86 | 59 | 41.03 |
| cohabitation | 1972 | 31.97 | 65.53 | 34.47 |
| widowed | 95 | 1.55 | 38.46 | 61.54 |
| divorced | 456 | 7.39 | 75.93 | 24.07 |
| **Educational status [$\chi^2 = 8.591$; $p < 0.035$]** |  |  |  |  |
| no formal education | 683 | 11.07 | 63 | 37 |
| primary education | 3984 | 64.6 | 66 | 34.11 |
| secondary education | 1231 | 19.96 | 64.06 | 35.94 |
| higher education | 269 | 4.37 | 52.27 | 47.73 |
| **Religion [$\chi^2 = 2.145$; $p = 0.543$]** |  |  |  |  |
| Christianity | 5,981 | 96.99 | 63.37 | 36.63 |
| Islam | 113 | 1.83 | 68.97 | 31.03 |
| Traditional/animalist | 1 | 0.01 | 100 | 0 |
| no religion | 58 | 0.95 | 60 | 40 |
| Other | 13 | 0.21 | 0 | 100 |
| **Parity [$\chi^2 = 22.550$; $p < 0.0001$]** |  |  |  |  |
| 1 - 3 | 4,026 | 65.28 | 67.19 | 32.81 |
| 4 - 6 | 1,667 | 27.03 | 50.49 | 49.51 |
| 7 - 9 | 423 | 6.86 | 54 | 46 |
| 10 or more | 51 | 0.83 | 100 | 0 |
| **Occupation [$\chi^2 = 11.190$; $p < 0.048$]** |  |  |  |  |
| did not work | 1,044 | 16.94 | 56.34 | 43.66 |
| Professional/technical/managerial | 173 | 2.8 | 56.63 | 43.37 |
| clerical | 37 | 0.6 | 59 | 41 |
| agriculture | 2,487 | 40.32 | 68.39 | 31.61 |
| household and domestic service | 823 | 13.35 | 62.36 | 37.64 |
| Others | 1,603 | 25.99 | 67.18 | 32.82 |
| **Wealth index [ $\chi^2 = 11.876$ ; $p < 0.018$ ]** |  |  |  |  |
| Poorest | 1,417 | 22.99 | 69.23 | 30.77 |
| Poorer | 1,191 | 19.31 | 65.97 | 34.03 |
| Middle | 1,198 | 19.42 | 69.74 | 30.26 |
| Richer | 1,208 | 19.59 | 62.63 | 37.37 |
| Richest | 1,153 | 18.69 | 56.93 | 43.07 |
| **Place of residence [ $\chi^2 = 15.170$ ; $p < 0.0001$ ]** |  |  |  |  |
| Urban | 1,099 | 17.82 | 55.0 | 45.0 |
| Rural | 5,068 | 82.18 | 67.5 | 32.5 |
| **Frequency of reading newspaper/magazine [ $\chi^2 = 1.680$ ; $p < 0.195$ ]** |  |  |  |  |
| not at all | 5,797 | 94.01 | 64.19 | 35.81 |
| at least once a week | 370 | 5.99 | 58.27 | 41.73 |
| **Frequency of listening to radio [ $\chi^2 = 2.677$ ; $p < 0.102$ ]** |  |  |  |  |
| not at all | 2,593 | 42.05 | 67.0 | 33.0 |
| at least once a week | 3,574 | 57.95 | 61.71 | 38.29 |
| **Frequency of watching television [ $\chi^2 = 17.255$ ; $p < 0.0001$ ]** |  |  |  |  |
| not at all | 5,158 | 83.64 | 67.40 | 32.60 |
| at least once a week | 1,009 | 16.36 | 53.47 | 46.53 |
| **Gestational age at ANC registration (months) [ $\chi^2 = 3.458$ ; $p < 0.484$ ]** |  |  |  |  |
| No ANC | 139 | 2.26 | 50 | 50 |
| < 4 | 3617 | 58.65 | 61.95 | 38.05 |
| 4 - 5 | 1650 | 26.76 | 67.54 | 32.46 |
| 6 - 7 | 636 | 10.32 | 66.67 | 33.33 |
| 8 or months | 121 | 1.97 | 50 | 50 |
| Don't know | 3 | 0.05 | 100 | 0 |
| **Number of ANC visits [ $\chi^2 = 13.350$ ; $p < 0.004$ ]** |  |  |  |  |
| None | 139.2283364 | 2.26 | 50.00 | 50.00 |
| < 4 | 3,115.80 | 50.52 | 67.34 | 32.66 |
| 4 - 8 | 2,901.24 | 47.04 | 61.00 | 39.00 |
| 9+ | 10.7365816 | 0.17 | 0.00 | 100.00 |
| **Person providing assistance during ANC [ $\chi^2 = 28.952$ ; $p < 0.0001$ ]** |  |  |  |  |
| Doctor | 278.6886712 | 4.52 | 44.6 | 55.4 |
| Nurse/midwife | 5,728.35 | 92.89 | 66.67 | 33.33 |
| Other health worker | 20.7318366 | 0.34 | 0 | 100 |
| No ANC | 139.2283364 | 2.26 | 50 | 50 |
| **Received 2+ tetanus injections [ $\chi^2 = 4.583$ ; $p < 0.032$ ]** |  |  |  |  |
| No | 4,095.74 | 66.41 | 60.94 | 39.06 |
| Yes | 2,071.26 | 33.59 | 67.63 | 32.37 |
| **Took iron tablet/syrup during pregnancy [ $\chi^2 = 0.028$ ; $p < 0.866$ ]** |  |  |  |  |
| No | 1,194.75 | 19.37 | 64.05 | 35.95 |
| yes | 4,972.25 | 80.63 | 63.34 | 36.66 |
| **Took intestinal parasite drugs during pregnancy [ $\chi^2 = 0.003$ ; $p = 0.958$ ]** |  |  |  |  |
| No | 3,515.91 | 57.01 | 64 | 36 |
| yes | 2,651.09 | 42.99 | 63.36 | 36.64 |

### Backward Stepwise Regression and Fitness of model

The original regression model used for the backward stepwise regression model included age, marital status, educational status, Parity, Occupation, Wealth index, place of residence, frequency of reading newspaper/magazine, frequency of listening to radio, frequency of watching, number of ANC visits, person providing assistance during ANC, and received 2+ tetanus injections. The final model, after removing variables that were non-significant at p-value ≤ 0.25, used for the regression analysis included age, occupation, place of residence, frequency of watching television, person providing assistance during ANC, and received 2+ tetanus injections.

The multicollinearity test showed that there was no evidence of collinearity among the independent variables (mean VIF = 1.14, minimum VIF = 1.03 and maximum VIF = 1.33). Also, Hosmer-Lemeshow test of the model was better explained by the model (chi-square 13.26, with 8 degrees of freedom and p-value = 0.1033). The AIC of the final model had a slightly lower value (1281.206) indicating a better model parsimony. The LRT showed non-significance p-value (p = 0.625) suggesting a similar a better fit to the data with the fewer number of predictor variables compared to the original model (reduced from 10 to 6) (table 2).

**Table 2:** Backward Stepwise Regression and goodness of fit of data.

| variables | Original model |  | Final model |  |
| --- | --- | --- | --- | --- |
|  | OR (95%CI) | p-value | OR (95%CI) | p-value |
| Age | 1.515089 (1.255 - 1.828) | < 0.0001 | 1.582 (1.338 - 1.871) | < 0.0001 |
| Marital | 1.055017 (0.901 - 1.235) | 0.505 | Excluded |  |
| Education | 1.029633 (0.837 - 1.267) | 0.782 | Excluded |  |
| Parity | 1.14874 (0.871 - 1.515) | 0.326 | Excluded |  |
| Occupation | 0.9543345 (0.896 - 1.016) | 0.145 | 0.952 (0.895 - 0.994) | 0.118 |
| Place of residence | 0.7363472 (0.536 - 1.012) | 0.059 | 0.73 (0.536 - 1.741) | 0.046 |
| Watching television | 1.241563 (0.874 - 1.764) | 0.227 | 1.253 (0.902 - 1.741) | 0.179 |
| Number of ANC visits | 1.163507 (0.889 - 1.522) | 0.269 | Excluded |  |
| Person providing assistance during ANC attendance | 0.7819763 (0.553 - 1.106) | 0.165 | 0.758 (0.543 - 1.058) | 0.104 |
| Receive 2+ tetanus injection | 0.8200604 (0.616 - 1.092) | 0.174 | 0.81 (0.612 - 1.071) | 0.139 |
| **Model significance (Prob > chi2)** | **<0.0001** |  | **<0.0001** |  |
| **Pseudo R2** | **0.0511** |  | **0.0492** |  |
| **AIC** | **1286.595** |  | **1281.206** |  |
| **LR p-value** | **0.625** |  |  |  |

OR: odds ratio, CI: Confidence interval, AIC: Akaike Information Criterion, LR: likelihood ratio

Table 3 illustrates that women aged 30 – 39 years were notably 3.5 times more inclined to opt for elective CS [cOR: 3.532, 95%CI: 2.248 – 5.550] compared to those aged 20 years or below in the binary regression. The level of significance of age as a risk factor for elective CS was maintained in the adjusted model [aOR: 3.130, 95%CI:1.969 - 4.978]. Women living in rural areas were less likely to opt for elective CS in the binary regression model [cOR: 0.587, 95%CI: 0.448 – 0.768], however the place of residence lost its significance when adjusted for confounders. Also, mothers who were provided with ANC assistance by nurse/midwife were 40% less likely [cOR: 0.403, 95%CI: 0.280 – 0.579] to undergo elective CS, compared to those provided assistance by doctors. When adjusted for confounders, it was still observed that the mothers who were provided ANC by nurses/midwife were still less likely to undergo elective CS [aOR: 0.529, 95%CI: 0.349 – 0.803] compared to those attended by doctors.

**Table 3:** Regression models on predictors of Caesarean section.

| Variables | Model I COR [95% CI] | Model II AOR[95% CI] |
| --- | --- | --- |
| **Age** |  |  |
| <20 | Ref | Ref |
| 20 - 29 | 1.580 [ 0.952 - 2.622] | 1.416 [0.846 - 2.369] |
| 30 - 39 | 3.532*** [2.248 - 5.550] | 3.130 [1.969 - 4.978]*** |
| 40 - 49 | 3.374*** [1.900 - 5.990] | 2.863 [1.581 - 5.186]** |
| **Occupation** |  |  |
| Not working | Ref | Ref |
| Professional/technical/managerial | 0.988 [0.592 - 1.649] | 0.562 [0.320 - 0.988]* |
| Clerical | 0.903 [0.331 - 2.463] | 0.576 [0.202 - 1.638] |
| Agriculture | 0.596** [0.417 - 0.852] | 0.684 [0.462 - 1.014] |
| Household and domestic service | 0.779 [0.417 - 0.852] | 0.715 [0.463 - 1.102] |
| Others | 0.630* [0.421 - 0.944] | 0.742 [0.482 - 1.143] |
| **Place of residence** |  |  |
| Urban | Ref | Ref |
| Rural | 0.587*** [0.448 - 0.768] | 0.808 [0.577 - 1.130] |
| **Frequency of watching television** |  |  |
| Not at all | Ref | Ref |
| At least once a week | 1.799*** [1.361 - 2.377] | 1.290 [0.912 - 1.826] |
| **Person providing assistance during ANC** |  |  |
| Doctor | Ref | Ref |
| Nurse/midwife | 0.403 [0.280 - 0.579] *** | 0.529 [0.349 - 0.803]** |
| Other health worker | 1 | 1 |
| No ANC | 0.805 [0.889 - 1.735] | 0.892 [0.112 - 7.092] |
| **Received 2+ tetanus injections** |  |  |
| No | Ref | Ref |
| Yes | 0.747 [0.572 - 0.976]* | 0.826 [0.622 - 1.096] |

cOR: crude odds ratio, aOR: adjusted odds ratio, CI: Confidence interval, \*p < 0.05,
\*\*p < 0.01, \*\*\*p < 0.001, Ref: Reference

## Discussion

The rate of CS has seen a significant increase across the globe, despite growing concerns about the negative effect CS. Emergency and elective CS delivery are two categories under which CS is conducted. In Rwanda, the rate of CS increased from 2.2 to 15.6% in the last two decades (13). This study findings also showed a very high rate of elective CS, 36.6% among mothers in Rwanda. This rate is among the highest in SSA (41), falling closely below South Africa, which has a rate of 39.7% (42). The high rate of elective CS could be attributed to the increasing misconception that CS is safer than the traditional vaginal delivery (43). Also, mothers with bad experience with vaginal delivery may opt in for elective CS even in the absence of any indications for CS. Many mothers opt for elective CS due to fear of labor pain and the desire to have more control over the birthing process (44).

To have valid and reliable predictors of elective CS, compared to other similar studies (41,43,45), this study used backward stepwise regression to select most influential predictors of elective CS in Rwanda. By eliminating the unnecessary predictors of elective CS in this current study, the model is easier to understand and interpret, particularly as the study findings could influence decision-making. The important predictor variables identified were tested for their multicollinearity using VIF. The VIF test was also done in the study by Mohammed et al., (2022) (45). There was no evidence of multicollinearity in this study data implying that each predictor provides unique information not influenced by the other factors. This clarity can be crucial in decision-making processes where understanding the impact of individual factors is necessary. Furthermore, the study used Hosmer-Lemeshow test, LRT and AIC to test the validity and reliability of the findings obtained from the use of the regression models. The outcome of these model diagnostics showed that our study models were very appropriate to predict the factors that has influence on the uptake of elective CS among mothers in Rwanda using the latest nationally representative survey, the RDHS (38–40).

One of the key predictors of elective CS identified in this study is maternal age during delivery. Maternal age plays a very crucial role in healthcare services, most especially when it has to do with decision taking (46). The study findings showed that mothers who were 30 years and above were more than twice likely to choose elective CS. This study findings correspond with similar studies conducted (13,41,43,45). Older mothers may choose elective CS because of their previous experience with childbirth, particularly through vaginal delivery. Also, multiparity, which poses threat to obstetric outcome, is somehow linked to maternal age (47,48). Hence, the decision to select elective CS in other to avoid any complications that may arise from spontaneous delivery.

Mothers who were working, particularly in agriculture were found to be almost 60% less likely to undertake elective CS compared to those who were not working. This findings corresponds with a previous study in Bangladesh (45) where mothers who had occupations were less likely to undertake CS, but is in contrast with a similar studies (43,49). Mothers in agriculture are predominantly common in rural settings where there is less access to healthcare facilities capable of providing CS. This could influence their low uptake of elective. However, in the adjusted model, there was a shift where mothers in the professional/managerial working category was rather found to have a lower risk of elective CS compared to mothers not working. Women in professional or managerial roles often have higher levels of education and access to information, which might lead them to be more aware of the potential risks and complications associated with elective surgical procedures like CS.

Our study findings also showed that mothers in the rural area were less likely to undertake elective CS, compared to mothers in urban. Urban areas typically have better healthcare infrastructure and more hospitals that are equipped to perform elective CS. This accessibility makes it easier for urban mothers to opt for such procedures compared to their rural counterparts who might have limited options.

Additionally, our study found that mothers who were exposed to watching television at least once a weak were almost 80% more likely to accept elective CS, compared to mothers with who did not watch television at all. Mothers who watch TV regularly may be exposed to medical programs, advertisements, and news that provide information about the availability and benefits of elective CS. This exposure can influence their perceptions and understanding, making them more open to considering and accepting elective CS as a viable option. A similar findings of exposure to visual media was found to influence uptake of elective CS (50).

Our study findings also found that person providing assistance during ANC was consistently a predictor of elective CS, with mothers who were attended by nurse/midwife being 40% less likely to accept elective CS compared to those attended by doctors. Midwives and nurses often emphasize natural birth processes and may have more time to invest in patient education and support, encouraging non-interventional approaches unless medically necessary. Doctors, especially in high-risk cases, may be quicker to recommend interventions, including elective cesarean sections, due to their training and a more conservative approach to managing potential liabilities and complications.

### Strength and Limitations of the Study

The study facilitates generalizability of the study findings because it used a nationally representative dataset from the Rwanda Demographic Health Survey. The study employed a rigorous data sensitivity test to ensure the validity and reliability of the findings. Additionally, the study used the most recent RDHS data, providing recent evidence regarding the subject of elective CS in Rwanda. However, the data employed cross-sectional design limiting its ability to establish causal relationship. Also, the study is prone to recall bias and misclassifications as with every other cross-sectional study.

## Conclusion and Recommendation

This study identified key predictors of a growing worry of elective CS through a valid and reliable modelled data from the 2019-2020 RDHS. The study findings has shown a very high rate of elective CS, among mothers in Rwanda. Predictors such as maternal age during delivery, the place of residence (urban), frequency of watching television, and person providing ANC services (doctors) were identified. These identified predictors must be deeply considered in developing important measures and policies directed at minimizing the unwarranted use of CS in Rwanda, which has detrimental impact on maternal and newborn outcome. Directed interventions on reducing the rate of elective CS in Rwanda should be a central focus towards reducing the overall rate of CS to align with the recommendation by WHO.

## Data Availability

The data for this study is available at https://www.dhsprogram.com/data/available-datasets.cfm.

## Acknowledgement

We wish to acknowledge the DHS program for providing the consent to using the RDHS dataset for this study.

## Ethical Consideration

The study used secondary data obtained from the MEASURE DHS programme. Therefore, ethical approval and participant consent were not necessary. However, permission and approval was obtained from the DHS programme to download and use the data from https://www.dhsprogram.com/

## Author Contributions

Conceptualization - MHK, TD; Methodology – MHK & KOA; Data Curation: MHK & APF; Formal analysis – MHK & TC; Writing of Original draft – MHK, TD & TC; Writing – review & editing – MHK & APF; Supervision & Validation – Damien Punguyire

